# Quantifying H5N1 outbreak potential and control effectiveness in high-risk agricultural populations

**DOI:** 10.1101/2025.07.22.25331905

**Authors:** Izel Avkan, Suzanne Gokool, Louise E. Smith, Genevieve Clapp, Rachel Cox, Amy C. Thomas, Ellen Brooks-Pollock

## Abstract

Avian influenza is a global public health threat. Since 2021, the ongoing H5N1 panzootic has brought a major shift in H5Nx epidemiology, including unprecedented spread, wide host range and lack of seasonality. Infections in marine mammals, wildlife and livestock have heightened concern for human-to-human transmission and pandemic potential. Contact tracing and self-isolation are used as public health measures in the UK to manage contacts of confirmed human cases of avian influenza. In this study, we aimed to estimate potential outbreak sizes and evaluate the effectiveness of contact tracing and self-isolation in managing community outbreaks of H5N1 following spillover from birds to people.

We characterised contact patterns from an underrepresented agricultural population at high risk of avian influenza exposure through contact with birds (Avian Contact Study). Informed by these realistic social contact data, we modelled outbreak sizes using a stochastic branching process model.

Most simulations resulted in small-scale outbreaks, ranging from 0 to 10 cases. When the basic reproduction number was 1.1, contact tracing and self-isolation reduced the average outbreak size from 41 cases (95% Confidence Interval (CI): 37-46 cases) to 7 cases (95% CI: 6- 8 cases), preventing, on average, 8 out of every 10 infections. However, controls became less effective in reducing the outbreak size when a higher proportion of cases were asymptomatic. Overall, our findings suggest that contact tracing and self-isolation can be effective at preventing zoonotic infections. Increasing awareness, encouraging self-isolation, and detecting asymptomatic cases through routine surveillance are important components of zoonotic infection containment strategies.

## Introduction

Highly Pathogenic Avian Influenza (HPAI) continues to pose a threat to both animal and human public health. It is a highly contagious viral disease that affects both domestic and wild birds, with occasional spillover to mammals, including humans (1,2). The H5N1 virus was first detected in southern China in 1996, causing large poultry outbreaks in Hong Kong in 1997. The virus re-emerged in 2003 spreading among wild birds across Asia, and later in Africa, Europe, and the Middle East (3). In October 2020, the reassortment of poultry and wild bird viruses led to the emergence of a novel H5N1 strain of clade 2.3.4.4b in the Netherlands and spread across Europe (4). Between 2022 and 2024, this H5N1 variant spread to mammals—including foxes, seals, sea lions, and mink (5). In March 2024, the virus was detected for the first time in dairy cows in the USA with widespread cattle-to-cattle transmission and spillover to cats and humans (5). In March 2025, it was detected for the first time in a sheep in the UK (6). Given the continued evolution of the virus and its increasing ability to infect mammals, there is a growing concern about its potential for human-to-human transmission.

Avian influenza is a viral infection that can cause mild to severe illness, including death in humans. Since 1997, a total of 998 human cases has been reported in more than 23 countries, with approximately half resulting in death (3,7). Common symptoms in humans include conjunctivitis (eye redness), fever, and respiratory symptoms such as cough, sore throat, and shortness of breath (8). However, there is a big variability in the proportion of asymptomatic infections and the factors influencing symptomatology are not well understood due to limited human cases (8,9).

The current absence of sustained human-to-human transmission categorises the situation as Phase 3 of the WHO pandemic phases (8,10,11). However, cases without zoonotic exposure and limited human-to-human transmission have been documented (12,13). Data from family clusters found evidence of human-to-human transmission with an estimated reproduction number of 1.14 (13). Although the current global public health risk is low, reassortment between avian and human seasonal influenza viruses could result in antigenic changes that facilitate human-to-human transmission, potentially transitioning the situation to the next phase (14). This emphasises the need for continuous surveillance and the development of effective control strategies.

Contact tracing is a key public health strategy used in early-stage pandemic response to control the spread of infectious diseases (15,16). Its success depends on factors such as a small proportion of asymptomatic cases, long incubation periods, prompt identification of contacts, and public willingness to share information (16). Currently, contact tracing and self- isolation are used by UK public health teams to manage contacts of confirmed human cases of avian influenza (17). Depending on the type of exposure, contacts are placed under either active or passive follow-up for 10 days after their last exposure. Active follow-up involves daily phone calls to monitor for symptoms, while passive follow-up involves providing guidance on what to do if symptoms develop. If a contact develops symptoms, they are assessed as a possible case and are advised to self-isolate while arrangements are made for clinical assessment and investigation.

The accuracy of models investigating the effectiveness of contact tracing relies on understanding contact networks, such as the number of contacts people have and with whom. Several factors influence contact networks, such as occupation, residential setting (urban or rural), and age (18,19). However, rural communities are often underrepresented in social contact surveys (20,21). Given the global epizootic of H5N1 avian influenza over the past 5 years, individuals working closely with birds are at increased risk of potential exposure, yet their contact patterns remain poorly understood. Understanding their contact patterns is crucial for assessing transmission dynamics.

In this paper, we used a novel dataset on contact patterns from an underrepresented UK agricultural population at risk of exposure to avian influenza through contact with birds. We applied a branching process model to simulate potential outbreak sizes for different scenarios and assessed the effectiveness of contact tracing and self-isolation to manage outbreaks of avian influenza among humans given known parameters for human-to-human transmission.

## Methods

### Data description

We used data from the Avian Contact Study—a questionnaire open to individuals aged 18 or over in the UK who have contact with domestic and/or wild birds to investigate the potential transmission dynamics of HPAI and inform public health policies (22). Although the questionnaire was open to people aged 18 and above, no individuals aged 18 responded, so our dataset includes only those aged 19 and over. Respondents generally had high levels of bird exposure: 20% reported flock sizes over 10000 birds, and more than 60% reported daily contact with birds (22). The questionnaire was launched in-person at the British Pig and Poultry Fair (15-16 May 2024) and subsequently circulated online via various poultry forums and networks by word-of-mouth (15 May to 31 October 2024). The questionnaire was divided into four sections:

Section A: “About you”

Section B: “Your contact with any type of domestic or wildlife birds” Section C: “Your contact with other people”

Section D: “Your awareness of avian influenza (bird flu)”

Details of the eligibility criteria, questionnaire format, patient and public involvement, and key results are described in Thomas AC, Gokool S, Clapp G et al. (2024) (22).

We extracted self-reported contact data with other people to inform branching process models. Direct contact was defined as either physical contact (such as a handshake, hug, kiss, or contact sport), conversational contact (exchanging at least a few words), or contact by distance (being within 2 metres of another person). Respondents were asked how many direct contacts they had in the last 24 hours, with the response options “1-5”, “6-10”, “11- 15”, “16-20”, and “More than 20”. Respondents then provided further details about each contact, such as relationship to the contact, age group of the contact, location, duration, and type of contact (physical, conversational, or by distance). Although the categorical response included an option of “More than 20 contacts”, respondents could only provide further information for up to twenty contacts in the following questions.

Additionally, we used contact data from CoMix, a large-scale contact survey started during the COVID-19 pandemic and conducted in several European countries, including the UK, as a comparator dataset from the general population (23). We took data relating to the final survey round collected post-pandemic in November 2022. The data processing for CoMix is described in the Supplementary Material.

### Data analysis

Since the number of reported contacts was a categorical variable, we counted the number of contacts listed by each participant to obtain an integer variable. This allowed calculation of summary statistics, degree distributions, and informed the branching process model. We used the ‘Relationship to the contact’ column to count the listed contacts, due to its lower proportion of missing data. We then compared the number of listed contacts to the reported contact category. No exclusions were made if the number of listed contacts did not align with the reported contact category. The number of listed contacts was used in our model.

We calculated the average number of contacts per person per day overall and stratified by age group and compared these values to CoMix (23). Distributions of contacts were calculated for three age groups (19-44, 45-59, 60+) representing young adults, adults, and older adults, as well as for all age groups combined. We fitted negative binomial and Poisson distributions to the data using the Maximum Likelihood Estimate method from the ‘fitdistrplus’ package version 1.2-1 in R and compared both fits using the likelihood ratio test.

Social networks commonly exhibit structural heterogeneity, and contact matrices are a widely used method for representing contact heterogeneities across age groups (24,25). We constructed age-stratified contact matrices to illustrate the average number of contacts made between different age groups, both overall and stratified for family contacts. We grouped ages into 10-year intervals to balance the level of detail without compromising sample sizes in each age group.

### Model framework

We used a stochastic branching process model to estimate the potential outbreak size and effectiveness of contact tracing and self-isolation in preventing the spread of avian influenza (Fig 1) (26–28).

**Fig 1.**
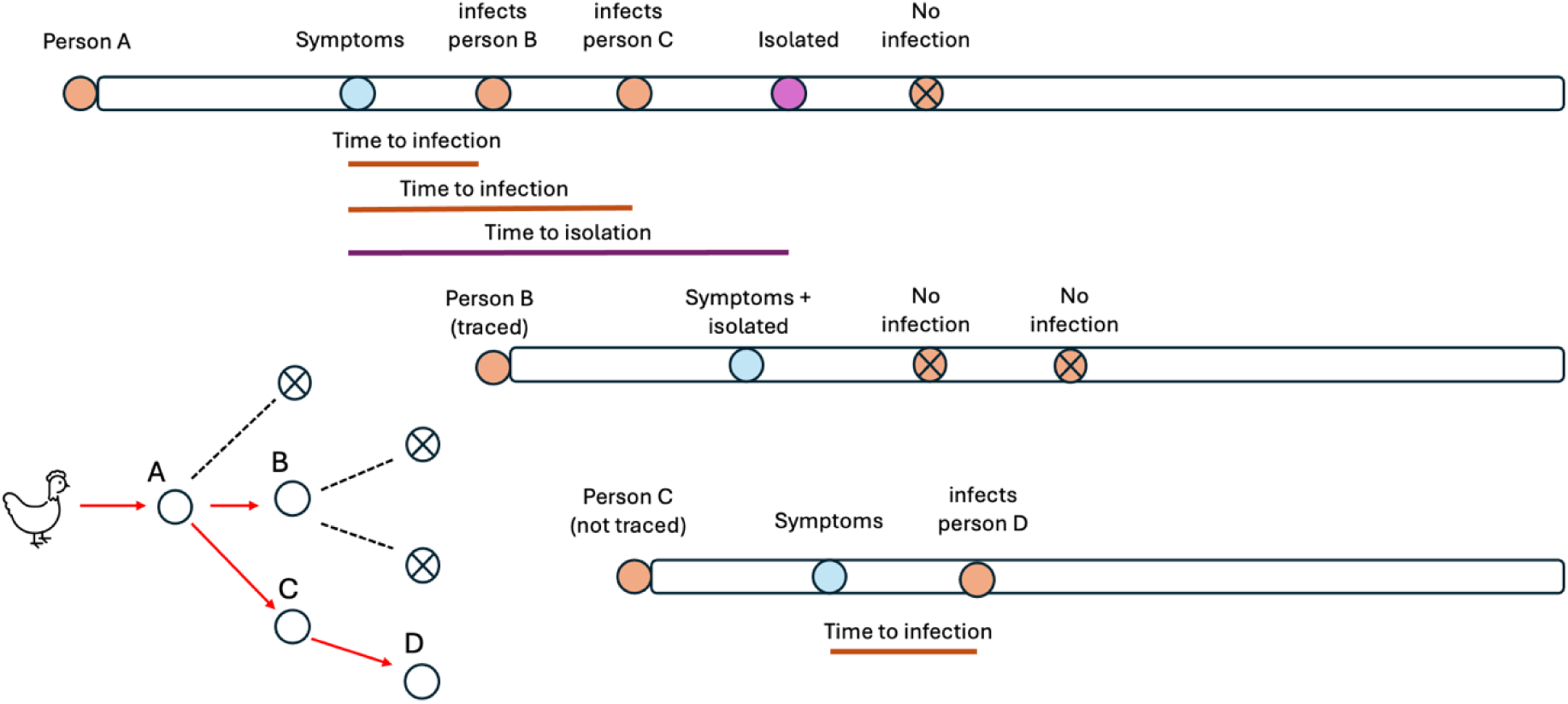
Contact Tracing Schematic. Example of a simulated transmission chain starting with person A, the index case, who was infected through contact with an infected bird. The number of secondary infections generated by each person is drawn from a negative binomial distribution. The time of infection for new cases is drawn from a gamma distribution, relative to the symptom onset of the infector. In this example, Person A infects 2 individuals, B and C, before isolating. If a contact is traced, they isolate on the day they develop symptoms and do not cause further infections. They may still be able to infect other people before showing symptoms. If a traced contact does not develop symptoms, or a contact is not traced, they would not isolate.

We defined outbreak size as the number of individuals in a cluster including the index case and analysed how it changed with the basic reproduction number, 𝑅_0_, the average number of secondary infections generated by an infected individual in a wholly susceptible population (29). We varied 𝑅_0_from 0 to 1.1 in increments of 0.1, consistent with estimates from previous studies on avian and pandemic influenza (13,30). To capture contact heterogeneities with age, we divided the population into four age groups (0-18, 19-44, 45-59, 60+). Since our dataset did not include individuals under 19 years old, we did not explore scenarios where index cases were under 19, but they are included in transmission chains using contact data from CoMix.

#### Number of secondary cases

The number of contacts made by an infected individual was drawn from an age-specific negative binomial distribution. The number of secondary infections (𝑅) generated by that case was calculated as the product of the number of contacts (𝑛), a fixed infectious period (𝑑), and the probability of transmission (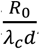, where 𝑅_0_ is the basic reproduction number, 𝜆𝑐 is the dominant eigenvalue of the contact matrix). Mathematically, the number of secondary infections generated by an individual in age group 𝑎 (for age groups 19-44 years, 45-59 years and 60+ years) is given by:

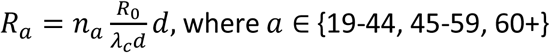

The age group of each secondary case was determined by sampling from a multinomial distribution, using probabilities from the corresponding row of the contact matrix, normalised to one. Time of infection for each secondary case relative to the symptom onset of the primary case was drawn from a shifted Gamma distribution taken to reproduce estimates from the literature (shape parameter 13, rate parameter 2, resulting in a mean of 3.5 days (SD: 1.80 days) between the symptom onset and infection) (8,13,29–32). We adjusted the distribution to allow infections to happen up to one day prior to the onset of symptoms.

#### Asymptomatic infections

The probability of being asymptomatic may depend on factors such as age and the strain of the virus. However, due to limited data on human cases of H5N1 avian influenza, the proportion of asymptomatic cases and the impact of age on this probability are not well understood. For the baseline scenario, we assumed 40% of individuals developed asymptomatic infections (8,9). However, as the proportion of asymptomatic infections has a major impact on the effectiveness of contact tracing and self-isolation, we explored a full range of scenarios from 0% of infections being asymptomatic to 100%

#### Contact tracing and self-isolation

We investigated the impact of contact tracing and self-isolation on the outbreak size. We assumed that 40% of contacts were traced in the baseline scenario, based on an estimate from reported figures during the COVID-19 pandemic (33). We determined whether new cases would be traced based on their symptom status. By default, the index case and the contacts of asymptomatic cases were not traced. Contacts of symptomatic cases could be traced with probabilities varying from 0 (no contact traced) to 1 (all contacts traced). For the purposes of this model, we defined self-isolation as the separation from others of symptomatic individuals who had been traced by public health teams, beginning on the day of symptom onset, in accordance with current guidelines. Traced contacts who do not develop symptoms, and untraced contacts, would not self-isolate.

We ran the model 9000 times per parameter set with the maximum time for each simulation of 100 days.

We used R version 4.4.3 in our analyses.

### Ethics Statement

Ethical approval for the study was obtained from the University of Bristol, Faculty of Health Sciences Research Ethics Committee, approval number 17048 on 16 January 2024.

Informed written consent (using e-consent hosted on REDCap) for the use of data collected via the questionnaire was obtained from respondents. A copy of the consent form and participant information sheet is given in the accompanying underlying data.

## Results

A total of 268 respondents completed the questionnaire between May and October 2024. We excluded 31 respondents who did not report a categorical number of contacts, and 40 who selected a category but did not provide further details, as we could not obtain an integer value from their responses. Among the 197 respondents who provided data on the categorical question about number of contacts and who listed individual contacts, 82% were consistent.

### Contact patterns

The distribution of numbers of contacts is shown in Fig 2A. The average number of contacts was 4.64 (standard deviation (SD): 3.40). A Negative Binomial (NB) model provided a better fit than a Poisson model based on Akaike Information Criterion (NB: 968, Poisson: 1052), Bayesian Information Criterion (NB: 974, Poisson: 1056), and likelihood ratio test (p-value < 0.001). The final NB model had a mean of 4.64 and a dispersion parameter of 4.02.

**Fig 2.**
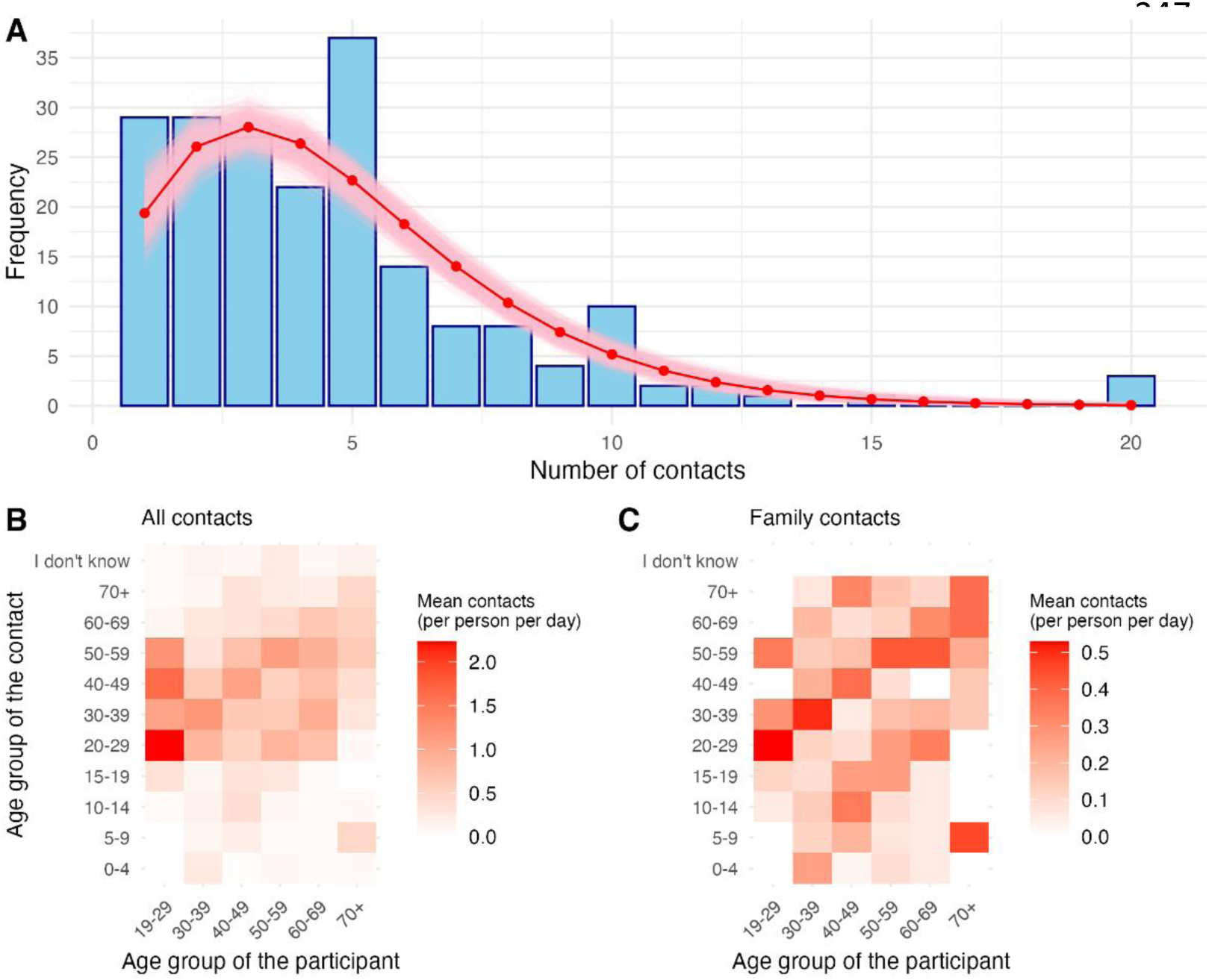
Contact patterns. Contact patterns reported in the Avian Contact Study. **(A)** the distribution of the number of contacts per person. The red line indicates the fitted negative binomial distribution, and the pink region represents the 95% confidence interval. The fitted negative binomial distribution had a mean of 4.64 and a dispersion parameter of 4.02. **(B)** Average number of contacts between age groups, while **(C)** Average number of contacts between family members only. In both **(B)** and **(C)**, darker colours indicate higher average contacts.

Additionally, we fitted NB distributions to the number of contacts stratified by age group to inform the branching process model, shown in S2 Fig.

### Contact matrices suggest intergenerational living in agricultural workers

We constructed age-structured contact matrices to show the average number of contacts made between different age groups, both for all contacts and family members (Fig 2B, C).

We observe broadly similar patterns to previous contact surveys, with respondents interacting more frequently with individuals of a similar age. Respondents aged 19-29 reported an average of 6.8 contacts per day, with nearly 90% of their contacts occurring with individuals aged 20-59. Respondents aged 30 and above had an average of 4.4 contacts per day.

Fig 2C illustrates contact patterns among family members. Respondents reported an average of 1.8 family contacts per day, with the 19-29 age group having the lowest (mean: 1.35 contacts, SD: 0.18 contacts) and the 40-49 age group the highest (mean: 1.97 contacts, SD: 0.14 contacts). We observed three distinct diagonal patterns: younger individuals frequently interacting with their parents, older individuals with their children and grandchildren, and individuals interacting with those of a similar age, reflecting intergenerational living arrangements.

A comparison of the Avian Contact Study and CoMix is shown in Fig 3. Respondents in the Avian Contact Study had higher average contact levels than respondents of CoMix across all age groups. The smaller sample size in the Avian Contact Study compared to CoMix may explain the wider confidence intervals around the mean. Statistical comparisons of the contact patterns are provided in the Supplementary Materials.

**Fig 3.**
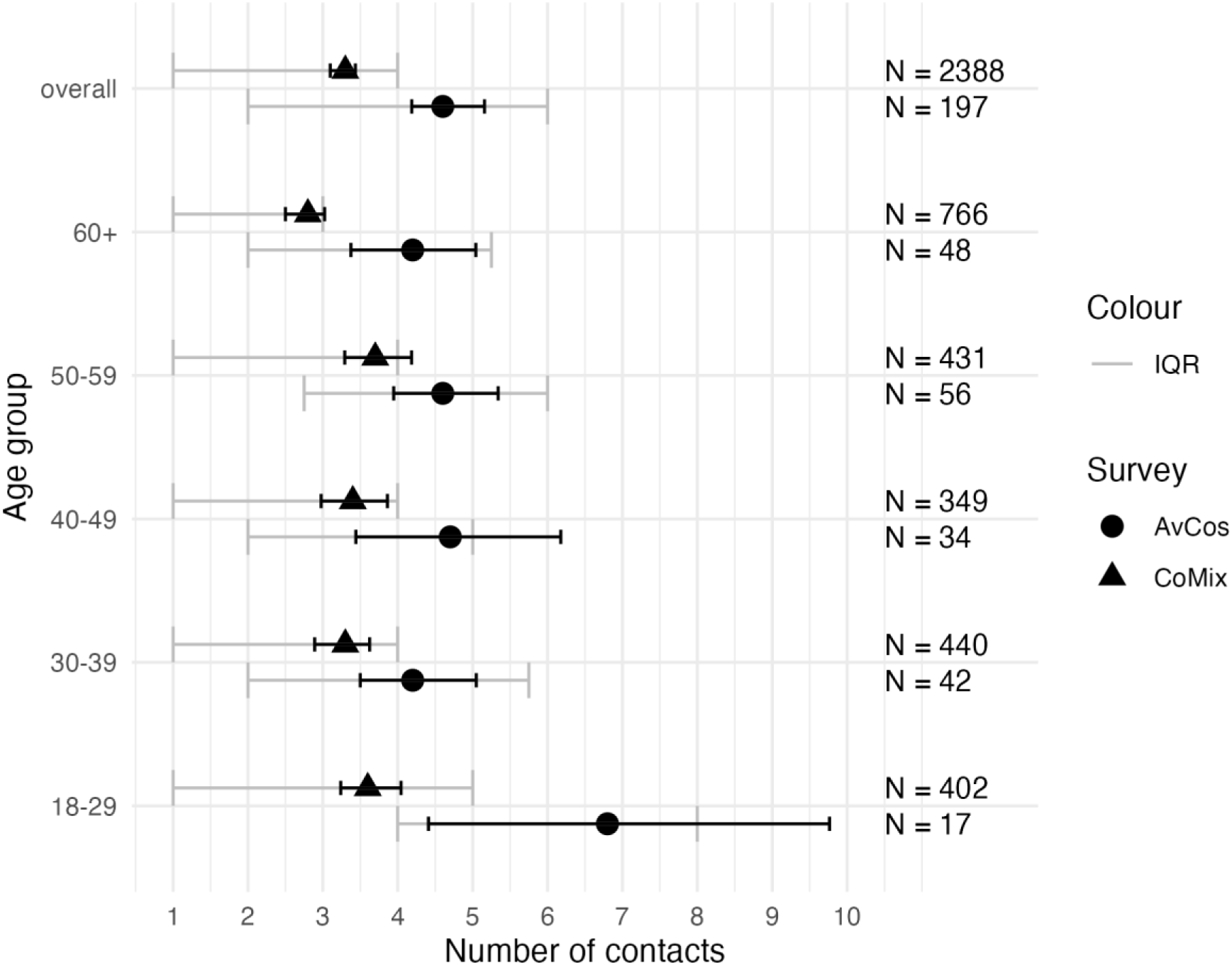
Average number of contacts from the Avian Contact Study (AvCos) and CoMix for all age groups. Black lines indicate the 95% confidence interval, and grey error bars indicate the interquartile range (IQR).

### Influence of age on outbreak size

As expected, outbreak size was highly dependent on the reproduction number. Infections with reproduction numbers less than 0.2 did not lead to onward transmission. Over 75% of simulations resulted in small-scale outbreaks (0-10 cases). The average outbreak size increased sharply from 4 cases at 𝑅_0_=1 to 41 cases at 𝑅_0_=1.1 (Fig S3).

We also examined how the age group of the index case influenced outbreak dynamics. For 𝑅_0_ between 0.7 and 1, half of simulations with index cases aged 60+ resulted in no secondary cases, compared to 40% of simulations for those aged 19-59. When 𝑅_0_ = 1.1, 9% of simulations with index cases in the 19-59 age group led to outbreaks larger than 50 cases, compared to 6% for those with index cases aged 60+.

For index cases in the 19-44 and 45-59 age groups, 𝑅_0_ did not significantly affect the age distribution of the cases. In contrast, for index cases aged 60+, we observed an increase in the percentage of cases in the 0-18 age group from 0 to 13% as 𝑅_0_increased. This is due to limited direct contact between the 60+ and 0-18 age groups, meaning individuals aged 0-18 were infected further down the transmission chain through interactions with other age groups (Figs S3 and S4).

### Reduced effectiveness of contact tracing and self-isolation with higher proportion of asymptomatic infections

We compared how outbreak sizes changed when 40% of infections were asymptomatic, under both no control and control (contact tracing and self-isolation) scenarios, depending on *R_0_* (Fig 4A). Under no control, no secondary cases were observed when 𝑅_0_was below 0.2 - the only case was the index case. Over 75% of simulations resulted in small-scale outbreaks (less than 10 cases). Contact tracing and self-isolation reduced the maximum outbreak size from 5705 cases to below 1000, and the average outbreak size from 41 cases to below 10, when 𝑅_0_=1.1.

**Fig 4.**
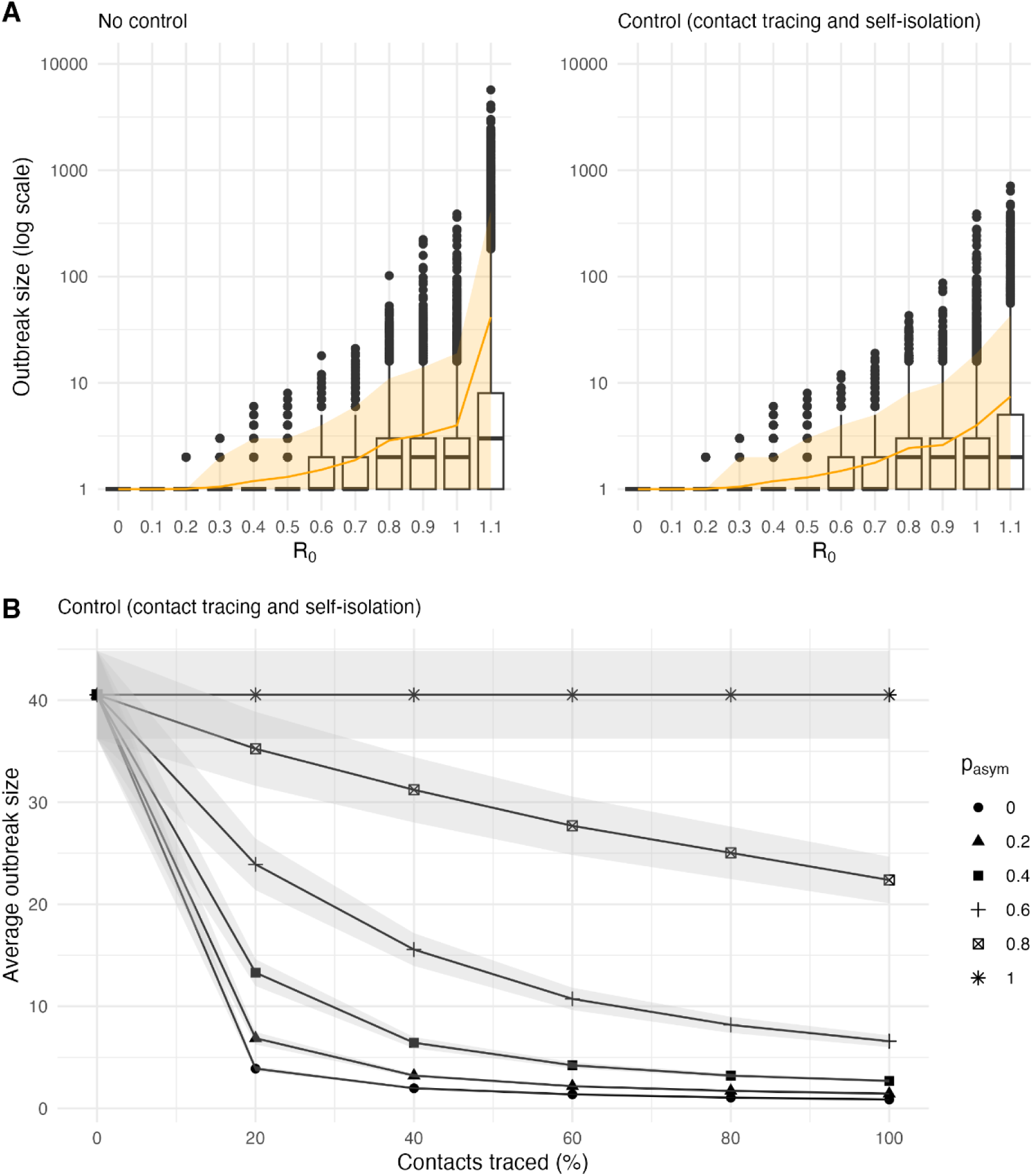
The impact of contact tracing and self-isolation on final outbreak size. **(A)** The distribution of outbreak sizes for varying levels of 𝑅_0_under no control and control (contact tracing and self-isolation). The orange line indicates the average outbreak size, and the shaded region represents the 95% prediction interval. We assumed that 40% of cases were asymptomatic and that 40% of contacts were traced. Among those traced, all symptomatic cases were assumed to self-isolate. **(B)** The average outbreak size across 9000 simulations for varying levels of contact tracing and proportions of asymptomatic infections when 𝑅_0_ = 1.1. Shaded regions represent the 95% confidence interval.

We also evaluated how the average outbreak size changed under different levels of control measures and varying proportions of asymptomatic infections (Fig 4B). When the proportion of asymptomatic infections was low, even low to moderate levels of contact tracing and self- isolation (0 to 40%) were sufficient to reduce the average outbreak size from 41 cases to below 10. However, the reduction in the average outbreak size became less pronounced as the proportion of asymptomatic infections increased.

## Discussion

### Principal findings

Our model provides insights into the potential outbreak size of avian influenza among humans under different levels of transmission. Most simulations resulted in small-scale outbreaks, with outbreaks exceeding 100 cases when the basic reproduction number was above 0.8, consistent with previous estimates (34). Contact tracing and self-isolation reduced outbreak size, but their potential effectiveness in preventing cases decreased as the proportion of asymptomatic infections increased, since they rely on symptomatic cases.

Previous modelling studies similarly concluded that contact tracing and self-isolation alone was highly unlikely to prevent large outbreaks, particularly when subclinical infections were present (28,35). Our findings support and extend these conclusions to a different pathogen.

### Translating findings into action

Asymptomatic infections limit the effectiveness of contact tracing and self-isolation, and they are optimally identified and controlled through asymptomatic testing programmes, which have previously shown to be successful at detecting cases (36). Reintroducing such programmes could enhance current interventions by improving detection of asymptomatic infections.

Moreover, we observed that the index cases aged 19-59 play an important role in transmission dynamics, leading to larger outbreaks based solely on contact data.

Additionally, analysis of previous H5N1 infections suggests that the infection is more prevalent among younger adults, and older age may not be a risk factor for severe outcomes (37). Taken together, these findings suggest that younger adults may benefit more from targeted interventions.

### Strengths and limitations

We used a novel dataset from the Avian Contact Study, which focused on individuals at higher risk of avian influenza and was developed with the help of public contributors to ensure relevance and accessibility (22). To our knowledge, it is the only contact survey conducted in the UK that has focused on farming communities since the COVID-19 pandemic. This allowed us to examine post-pandemic contact patterns in a rural, high-risk groups, often underrepresented in contact surveys, who may have different mixing patterns compared to the general population. We used age-stratified mixing patterns in our model to estimate outbreak sizes, extending previous models that did not incorporate age structure. This allowed us to capture variation in outbreak sizes across age groups and identify which groups were more likely to drive larger outbreaks.

In addition to common limitations of other contact surveys (21,38), our data did not include individuals under the age of 19, so we could not compare their contact patterns to those of the general population. Although we included 0-18 age group from CoMix in the transmission chain, this may not fully reflect the contact patterns due to differences in the cohorts and study periods. Additionally, our analysis was based on data from individuals living in the UK, most of whom were poultry farmers, hence our findings may not be generalised to other countries or to farming communities with different livestock practices.

Although we included age in our model, we could not explore its relationship with all the parameters. For instance, the influence of age on symptomatology is not well understood. As a result, we kept the proportion of asymptomatic infections constant across all age groups. Our model assumed that people with symptoms would isolate on the same day, although this may not be the case in practice with people adopting a ‘wait and see’ approach (39).

### Areas for future work

Future research is required to further characterise contact patterns within agricultural demographics and other related groups. In addition to farmers who make up 60% of the agricultural workforce in the UK, 12% is covered by casual workers (40). Despite their high exposure risk, it can be challenging to engage with this group due to language and trust barriers.

## Data Availability

Data and code availability
Underlying data
Repository data.bris: The Avian Contact Study: questionnaire data 15 May to 31 July 2024.
Data are openly available at the University of Bristol Research Data. Repository data.bris, at https://doi.org/10.5523/bris.3nmqsrbv5ruom2abn0ql6e8yh2.
This project contains the following underlying data:
Data file 1. (Raw underlying questionnaire data, csv file)
Data file 2. (Raw underlying questionnaire data, .RDS file)
Data file 3. (Associated data dictionary, csv file)
Data file 4. (Code for importing underlying data in csv format into R for setting up labelled data .r file)
Data file 5. (Blank consent form and participant information sheet, pdf file)
Data are available under the terms of National Archives Non Commercial Government Licence for public sector information.
Extended data
Repository Zenodo: The Avian Contact Study Questionnaire and Data Dictionary [10.5281/zenodo.13617061]
This project contains the following extended data:
AvianInfluenzaSocialContactSu.pdf (The final questionnaire REDCap PDF)
AvianInfluenzaSocialContactSur_DataDictionary_080824v1.csv (Associated data dictionary csv file)
Data are available under the terms of the Creative Commons Attribution 4.0 International license (CC BY 4.0).
Software availability
Source code available from: https://github.com/amythomas/aviancontactstudy
License: Creative Commons Attribution 4.0 International license (CC BY 4.0).

## Acknowledgements

We are grateful to all respondents who completed the questionnaire and to the public contributors for their input into study design and questionnaire development. We thank all members of the wider Avian Contact Study team for input into study design, data collection and interpretation. We thank Dr Anastasia Chatzilena (University of Bristol) for her guidance regarding the statistical analysis of contact data from the Avian Contact Study and CoMix.

## Data and code availability Underlying data

Repository data.bris: The Avian Contact Study: questionnaire data 15 May – 31 July 2024. Data are openly available at the University of Bristol Research Data Repository <u>data.bris</u>, at https://doi.org/10.5523/bris.3nmqsrbv5ruom2abn0ql6e8yh2 (41).

This project contains the following underlying data:

- Data file 1. (Raw underlying questionnaire data – csv file)

- Data file 2. (Raw underlying questionnaire data - .RDS file)

- Data file 3. (Associated data dictionary – csv file)

- Data file 4. (Code for importing underlying data in csv format into R for setting up labelled data - .r file)

- Data file 5. (Blank consent form and participant information sheet – pdf file) Data are available under the terms of National Archives’ <u>Non-Commercial</u> <u>Government Licence for public sector information</u>.

## Extended data

Repository Zenodo: The Avian Contact Study Questionnaire and Data Dictionary [10.5281/zenodo.13617061]

This project contains the following extended data:

- AvianInfluenzaSocialContactSu.pdf (The final questionnaire REDCap – PDF)

- AvianInfluenzaSocialContactSur_DataDictionary_080824v1.csv (Associated data dictionary -csv file)

Data are available under the terms of the Creative Commons Attribution 4.0International license (CC-BY 4.0).

## Software availability

Source code available from: https://github.com/amythomas/aviancontactstudy

License: Creative Commons Attribution 4.0 International license (CC-BY 4.0).

