## Supplementary Figures for "Quantifying H5N1 outbreak potential and control effectiveness in high-risk agricultural populations"

**Supplementary Material**

**Pre-processing of CoMix data**

We used CoMix data to compare contact patterns with those from the Avian Contact Study, which is publicly available [here](https://zenodo.org/records/11154066). Specifically, we used the “participant_common” and “contact_common” datasets to obtain the number of contacts reported by each participant. The “participant_common” dataset includes information such as participant id, age group, and gender, while the “contact_common” dataset is in wide format and contains participant id, contact id, and details about each contact.

We extracted UK data, and included only participants aged above 18, as the Avian Contact Study recruited individuals over 18 living in the UK. We calculated the total number of contacts for each participant by counting the appearance of each unique participant id in the “contact_common” data. Note that participants with zero contacts do not appear in this dataset. We identified these participants from “participant_common” and assigned them a contact count of zero. Age group of the participants was also obtained from the “participant_common” dataset.

For consistency with the Avian Contact Study, we applied a maximum cut-off of 20 contacts per participant and used those contact counts in our analysis.

**Comparing contact data from the Avian Contact Study to CoMix**

We applied various methods to compare the contact patterns between the Avian Contact Study and CoMix. We provided a visual comparison of the probability distributions of the contact patterns in supplementary S1 Fig. The CoMix dataset showed higher probabilities for 0 to 3 contacts, while the Avian Contact Study showed higher probabilities for 4 to 12 contacts.

We also used statistical tests to compare the patterns, such as a chi-square test on grouped contact counts. We grouped the number of contacts into the following categories: 0-1, 2, 3, 4, 5, 6, 7-9, 10-20. This ensured that the expected frequency for each category is at least 5, which is one of the assumptions of the chi-square test. We also calculated the Total Variation Distance (TVD), defined as $\frac{1}{2}\sum|P\left( x \right)-Q\left( x \right)|$, where $P\left( x \right)$and $Q\left( x \right)$ are the probability distributions of the Avian Contact Study and CoMix, respectively. We then used a permutation test to assess the statistical significance of the estimated TVD by randomly shuffling the survey labels 10000 times. This produced a distribution of TVD values that would be expected if there was no difference between the groups. The p-value was then calculated by determining the proportion of TVD values in the permutation distribution that are greater than or equal to the observed value. We estimated the TVD to be 0.31, and the p-values from both the chi-square test and the permutation test less than 0.0001.

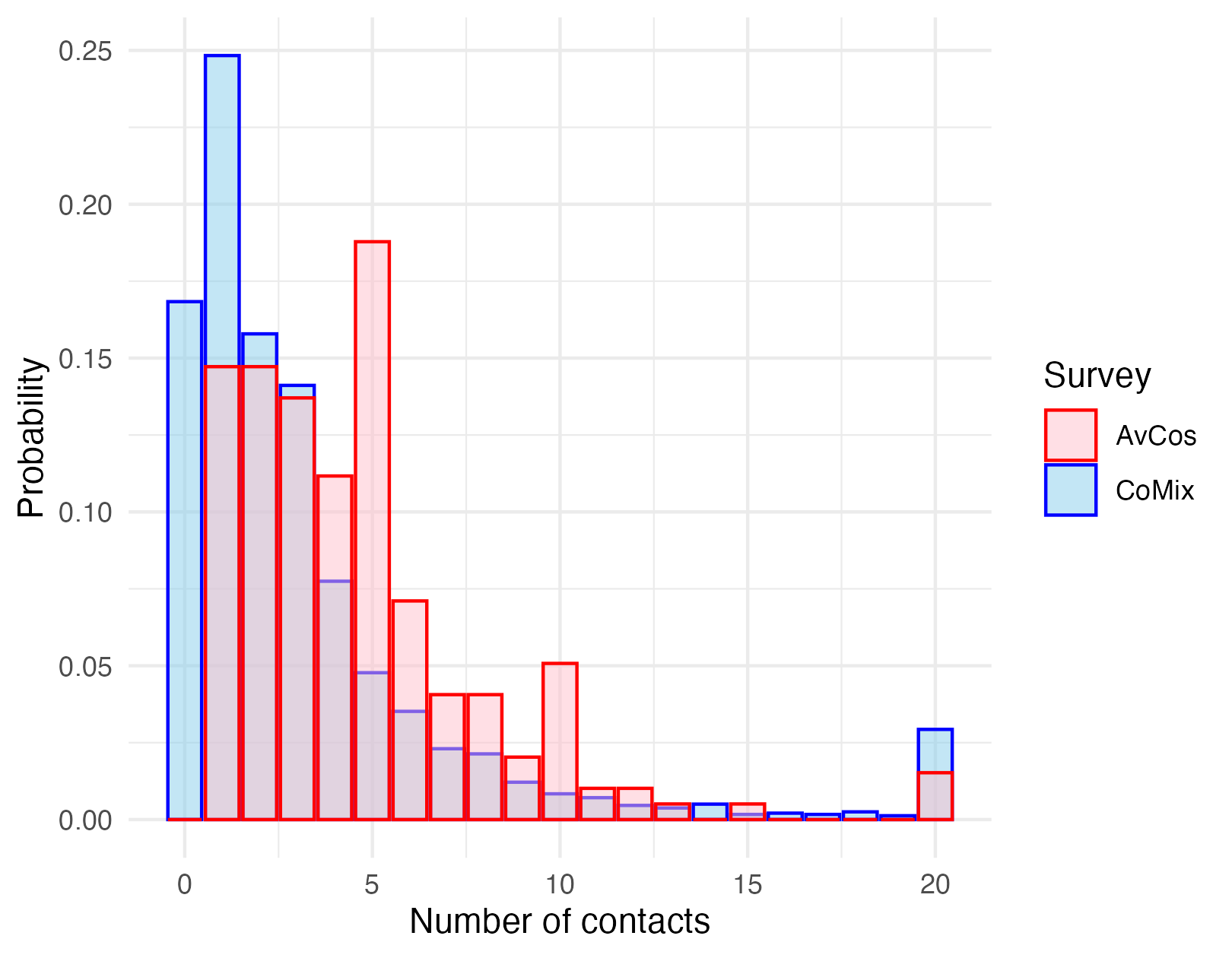
Overall, these findings suggest a statistically significant moderate difference in contact distributions between the two datasets. These differences may be due to differences in the settings of both surveys, such as the study periods and the target population.

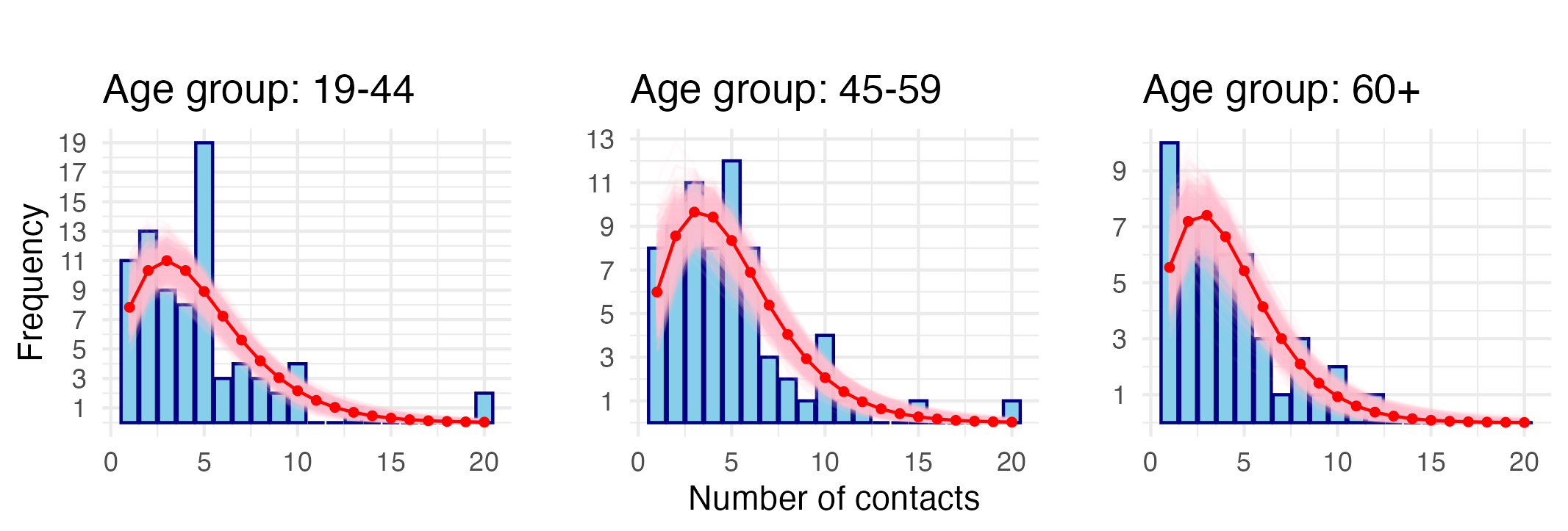
**S1 Fig.** **Comparison of the empirical probability distributions of the number of contacts reported in the Avian Contact Study (AvCos) and CoMix.**

**S2 Fig.** **Distribution of age-stratified number of contacts from the Avian Contact Study**. The red line indicates the fitted negative binomial distribution, and the pink region represents the 95% confidence interval.

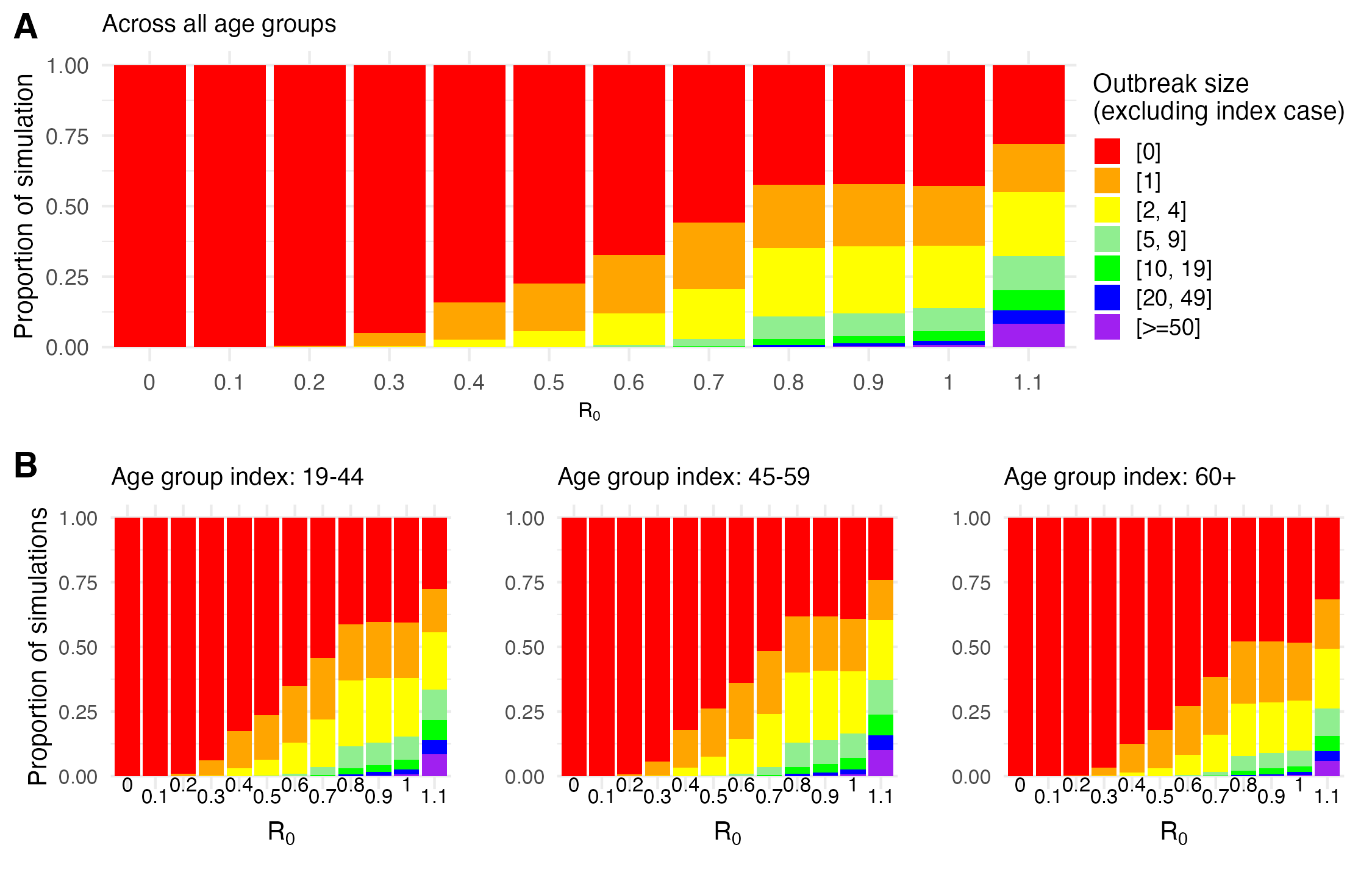

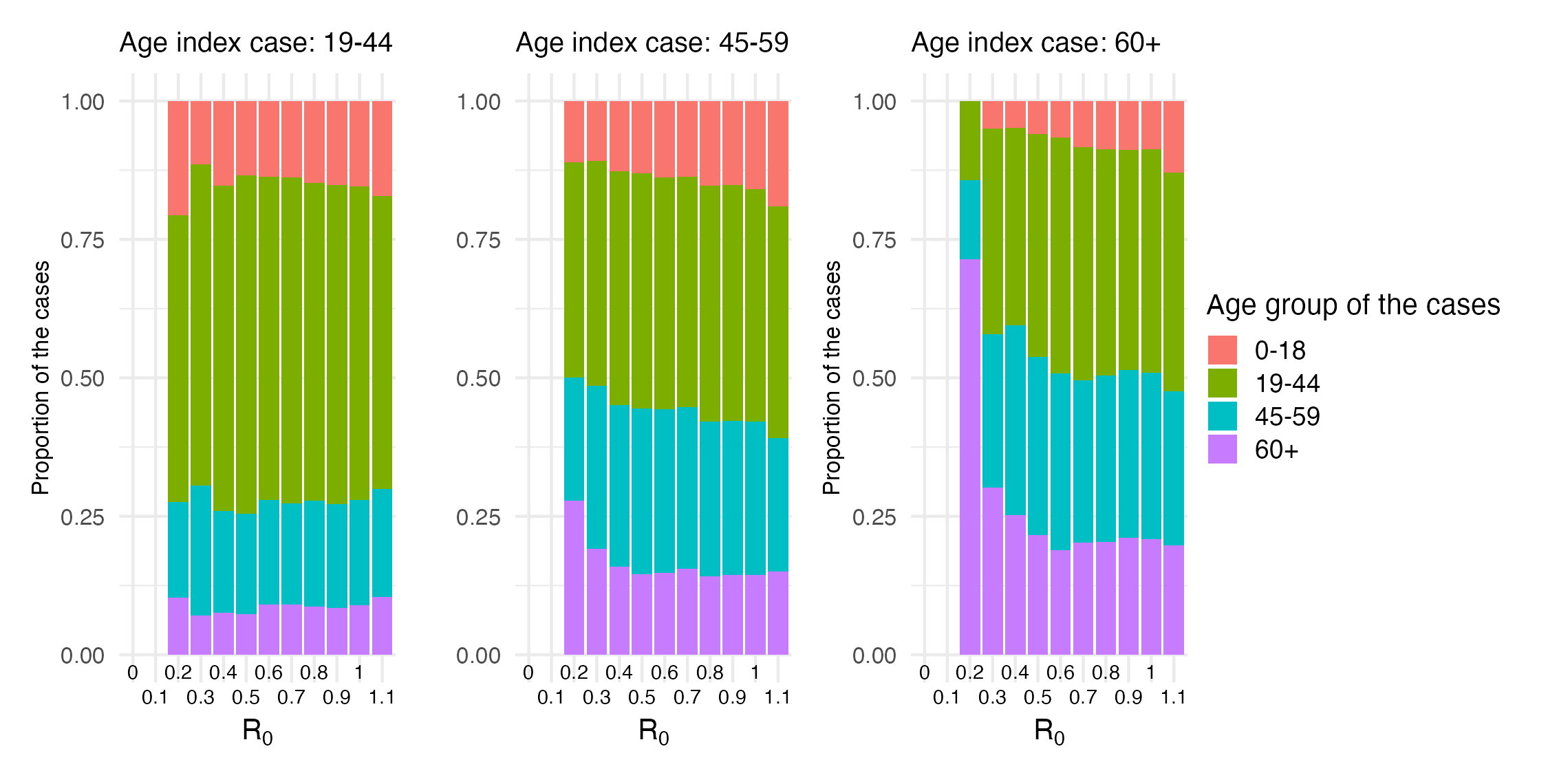
**S3 Fig.** **Effect of age and basic reproduction number on final outbreak size.** **(A)** Outbreak size across all age group. **(B)** Age-specific outbreak size. Each coloured bar represents a different outbreak size (excluding the index case), and the height of each bar indicates the proportion of simulations resulting in the corresponding outbreak size. We assumed 40% of cases were asymptomatic.

**S4 Fig.** **Impact of age of the index case on the age distribution of cases in a cluster.** Each coloured bar represents a different age group, and the height indicates the average proportion of cases resulting in that age group across 9000 simulations. No bars are shown for R_0_ values of 0 and 0.1, as no secondary cases occurred at those values. We assumed 40% of cases were asymptomatic.
